# Evaluation of a fully automated high-throughput SARS-CoV-2 multiplex qPCR assay with build-in screening functionality for DelHV69/70- and N501Y variants such as B.1.1.7

**DOI:** 10.1101/2021.02.12.21251614

**Authors:** Dominik Nörz, Moritz Grunwald, Flaminia Olearo, Nicole Fischer, Martin Aepfelbacher, Susanne Pfefferle, Marc Lütgehetmann

**Affiliations:** University Medical Center Hamburg-Eppendorf (UKE), Institute of Medical Microbiology, Virology and Hygiene, Hamburg, Germany

## Abstract

1

**Background:** New SARS-CoV-2 variants with increased transmissibility, like B.1.1.7 from England or B1.351 from South Africa, have caused considerable concern worldwide. In order to contain the spread of these lineages, it is of utmost importance to have rapid, sensitive and high-throughput detection methods at hand.

**Methods:** Analytical sensitivity was assessed for both wild-type SARS-CoV-2 and B.1.1.7 lineage by serial dilution. A total of 141 clinical samples were subjected to the test and results compared to a commercial manual typing-PCR assay and NGS.

**Results:** The multiplex assay is highly sensitive for detection of SARS-CoV-2 RNA in clinical samples, with an LoD of 25.82 cp/ml (CI: 11.61 – 57.48). LoDs are slightly higher for the HV68/70 deletion (111.36 cp/ml; CI: 78.16 – 158.67) and the N501Y SNP (2548.04 cp/ml, CI: 1592.58 – 4076.73). A total of 141 clinical samples were tested with the assay, including 16 samples containing SARS-CoV-2 of the B.1.1.7 lineage. Three non-B.1.1.7 samples contained a HV69/70 deletion. All were correctly identified by the multiplex assay.

**Conclusion:** We describe here a highly sensitive, fully automated multiplex PCR assay for the simultaneous detection of del-HV69/70 and N501Y that can distinguish between lineages B.1.1.7 and B1.351. The assay allows for high-throughput screening for relevant variants in clinical samples prior to sequencing.

## 2 Introduction

A number of novel SARS-CoV-2 variants that emerged in fall 2020, some of which show increased transmissibility and possible immune escape, are increasingly attracting worldwide attention (1, 2).

The B.1.1.7 lineage (VOC202012/01, 501Y.V1) first emerged in southern England and is notable for an unusually high number of mutations with no direct common ancestor compared to previous sequences (3, 4). In particular, non-synonymous spike-gene mutations such as the N501Y SNP (single nucleotide polymorphism) may lead to changes in receptor binding properties (5). Another variant, B.1.351 (501Y.V2) first described in south Africa, is characterized by the same spike mutation, along with E484K and others (6). Both have largely replaced previously circulating lineages in their respective areas of origin, indicating improved host adaptation and immune evasion.

As a result of increased awareness in public health and science institutions, there has been a rapidly growing demand for whole genome sequencing worldwide in order to recognize and map the spread of these emerging variants. In this context, the speed and scalability of fully automated RT-PCR can be highly beneficial to pre-screen samples for relevant mutations. The aim of this study was to create and validate a high-throughput first-line screening assay with the build-in ability to discriminate between relevant SARS-CoV-2 variants such as B.1.1.7 and B.1.351 on a fully automated sample-to-result PCR-platform (7). Inhouse assays have previously been used successfully for SARS-CoV-2 detection and diagnostics with this system via its open mode (Cobas Omni Utility Channel) (8).

## 3 Multiplex assay setup

The basic rationale of the multiplex assay was to combine two highly sensitive diagnostic SARS-CoV-2 PCR assays with additional assays to simultaneously detect relevant mutations. The SC2-assay by the US CDC (N-gene, SC2-N) was modified to serve as Pan-SCoV2 target (9). A publicly available diagnostic S-gene assay by Zhen et al. serves as second target (SC2-S) (10), while also featuring a drop-out phenomenon in the presence of a HV69/70 deletion due to probe location, which is rescued by an additional Taqman-probe (S-Del) that allows differentiation for wild-type and del-HV69/70. An additional assay was designed (using Beacon designer and PrimerQuest software) and integrated into the multiplex to detect the N501Y SNP. See *Figure 1* for a schematic overview of the PCRs and their target regions.

**Figure 1:**
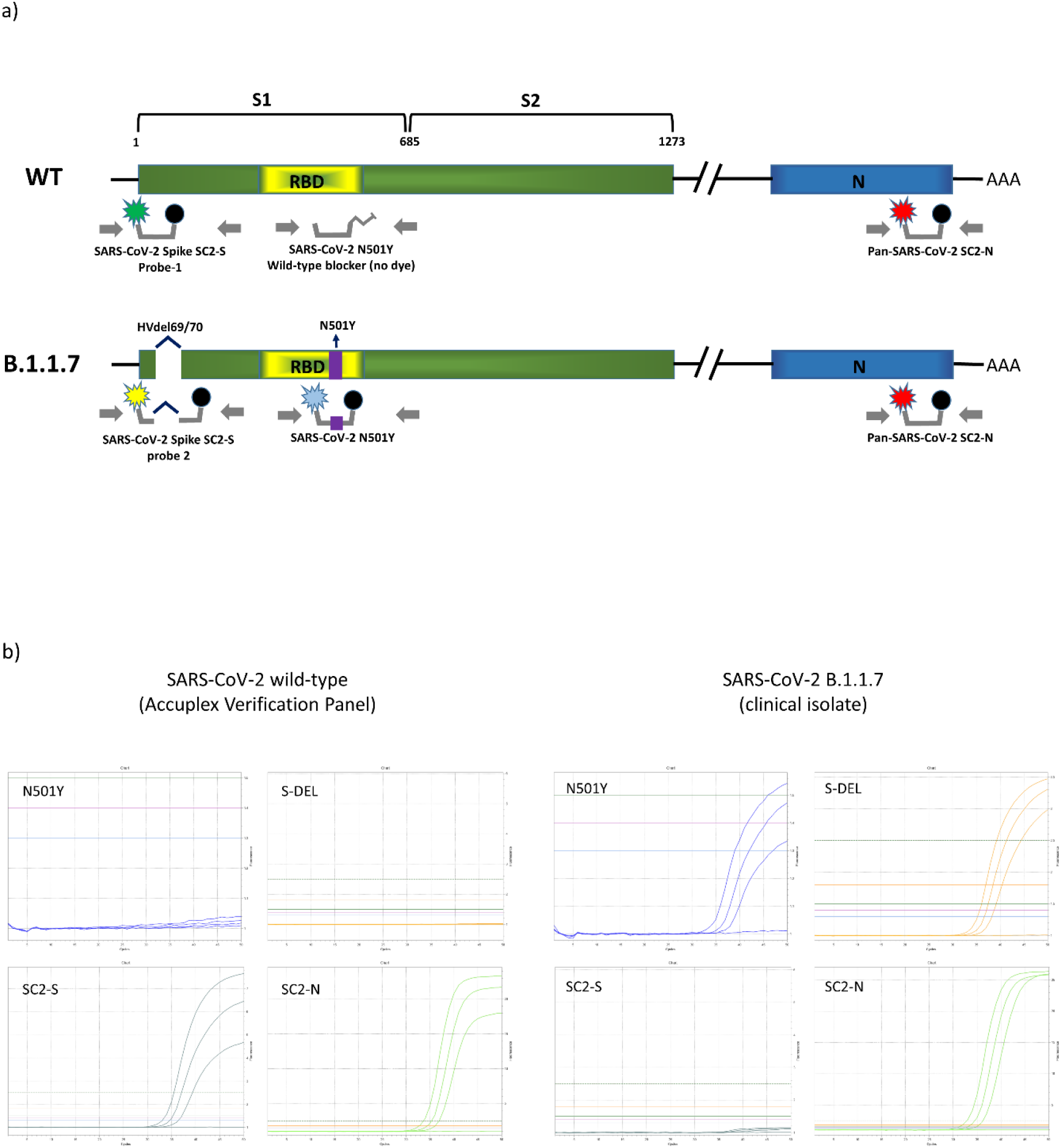
a) Schematic overview of the primers and probes used in the novel multiplex assay and their target regions within the SARS-CoV-2 lineages (wildtype (WT) and B.1.1.7). SARS-CoV-2 Spike SC2 PCR targets the spike gene (S1) of both lineages, with probe-1 detecting the wildtype sequence and probe 2 being specific for the HV69/70 deletion of the B.1.1.7 lineage. The SARS-CoV-2 N501YPCR targeting the receptor binding domain (RBD) is specifically designed for detecting the N501Y SNP. Both lineages are detected by the third PCR targeting the N-gene (N). b) Exemplary amplification curves for low concentration SARS-CoV-2 wild-type and B.1.1.7 as seen in the Utility Channel optimization software.

Primers and Probes were added to MMX-R2 to form the MMR2 mastermix and loaded into cobas omni utility channel cassettes, according to instructions by the manufacturer. Final concentrations of primers and probes are indicated in *table 1*. The utility channel run protocol is outlined in *table 2*.

**Table 1:**
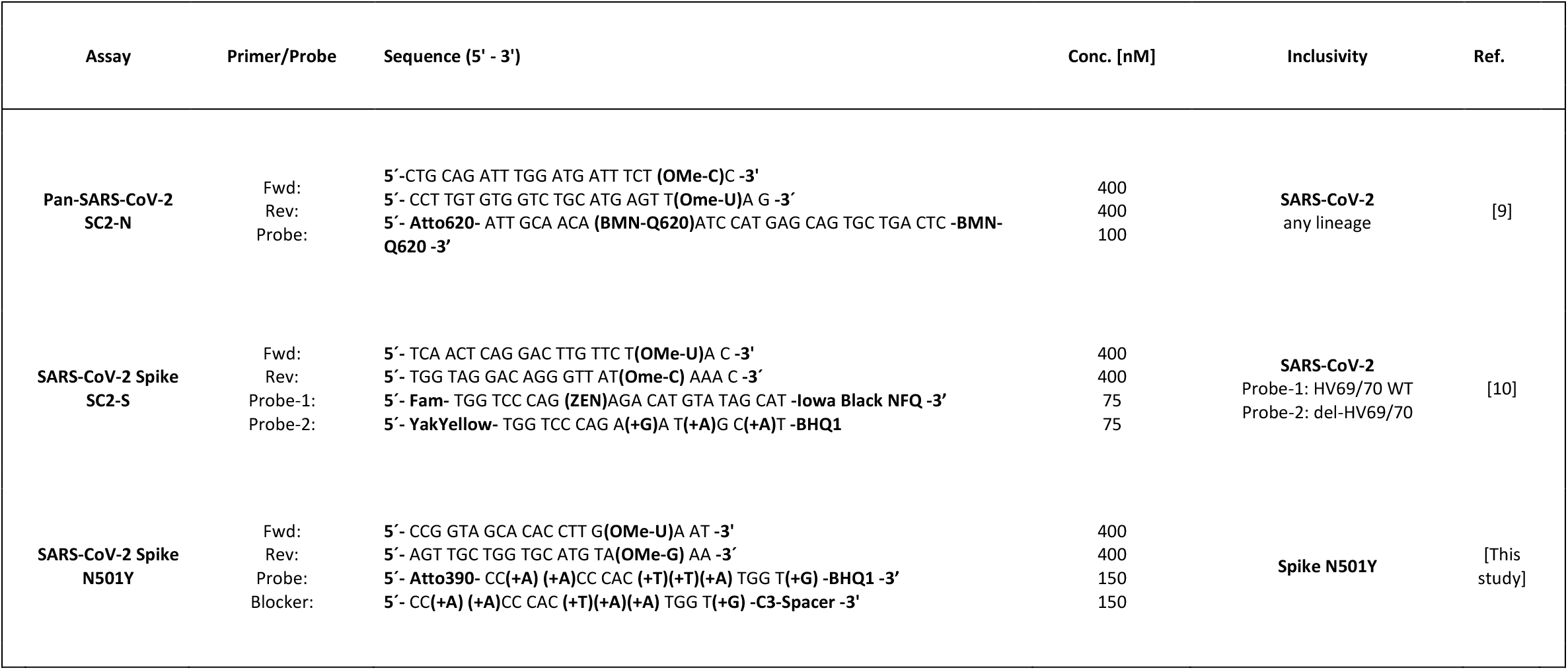
Primers and probes were custom made by Integrated DNA Technologies (Coralville, USA), Ella Biotech GmbH (Martinsried, Germany) and biomers.net GmbH (Ulm, Germany). Final concentrations of oligonucleotides indicated above refer to concentrations within the final reaction mix. OMe-X, 2’O-methyl-RNA base. BMN-Q620, proprietary dark-quencher by biomers.net can likely be replaced with e.g. BHQ2. +X, Locked nucleic acid (LNA) base.

**Table 2:** Run protocol for the SCOV2_VAR_UCT assay. Configuration was done with cobas omni Utility Channel software according to instructions by the manufacturer.

| Software settings |  |  |  |  |  |
| --- | --- | --- | --- | --- | --- |
| Sample type | Swab (400 µL) |  |  |  |  |
| Channels | 1: *N501Y* | 2: *SC2-S* | 3: *S-DEL* | 4: *SC2-N* | 5: *IC* |
| RFI | 1.3 | 1.5 | 1.8 | 1.4 | 1.5 |
| PCR cycling conditions |  |  |  |  |  |
|  | UNG incubation | Pre-PCR step | 1<sup>st</sup> measurement | 2<sup>nd</sup> measurement | Cooling |
| No. of cycles | Predefined | 1 | 5 | 45 | Predefined |
| No. of steps |  | 3 | 2 | 2 |  |
| Temperature |  | 55°C; 60°C; 65°C | 95°C; 55°C | 91°C; 58°C |  |
| Hold time |  | 120 s; 360 s; 240 s | 5 s; 30 s | 5 s; 25 s |  |
| Data acquisition |  | None | End of each cycle | End of each cycle |  |

The cobas 6800/8800 internal control (IC) is a spike-in (packaged) RNA target, which is automatically added during extraction by the system. MMRX-R2-reagent already contains the internal control assay by default; the respective sequences are not disclosed by the manufacturer. The IC acts as a full process control in the same way as in commercial cobas 6800/8800 IVD tests manufactured by Roche.

## 4 Evaluation of Sensitivity, Specificity and clinical performance

For analytical performance evaluation, quantified SARS-COV-2 reference material (Seracare Accuplex SARS-CoV-2, FluA/B and RSV Verification Panel, Milford, USA) and patient sample containing B.1.1.7 lineage (according to SARS CoV-2 whole genome sequencing) were used to prepare dilution series in pooled negative patient samples. Absolute quantification of the latter was carried out using the same assay with preexisting linearity data and the Accuplex verification panel as reference on the cobas6800 system (11). 2-fold dilution series (8 repeats per dilution step) were used to determine lower limit of detection.

To evaluate clinical performance and specificity of differentiation, a set of 101 samples RT-PCR positive for SARS-CoV-2 were subjected to the SCOV2_VAR_UCT. Of these samples, 14 were predetermined as B.1.1.7 lineage. Presence of the respective mutations was confirmed by a set of commercial assays by TIB MOL (VirSNiP Spike N501Y and Del69/70, Berlin, Germany). Presence of del-HV69/70 and N501Y mutations in the non-B117 set was precluded using the same commercial assays. A further 40 SARS-CoV-2 negative samples (by cobas SARS-CoV-2 IVD test), and a cross-reactivity panel containing various respiratory pathogens (*supplementary table 1*) were tested with the assay to assess overall specificity.

Overall analytic LoD (95% chance of detection) was determined as 25.82 cp/ml (CI: 57.48 – 11.61) (SC2-S LoD: 73.89; CI: 151-03 – 36.16. SC2-N LoD: 11.13, CI 16.02 – 7.73), as seen in *supplementary table 2*. There were no significant differences with B.1.1.7 lineage (Lod: 20.37, CI: 97.86 – 20.37) (S-DEL LoD: 111.36; CI: 158.67 – 78.16. SC2-N LoD: 20.37; CI: 97.86 – 20.37). Limit for successful detection of an N501Y SNP was 2548.04 cp/ml (CI: 4076.73 – 1592.58), indicating that the N501Y SNP-assay is about 100-fold less sensitive than the two diagnostic-grade SARS-CoV-2 assays.

Of the 101 SARS-CoV-2 positive samples, all were correctly identified by the SCOV2_VAR_UCT. 16 B.1.1.7 lineage samples were classified as positive for N501Y and del-HV69/70. 3 of the 85 non-B.1.1.7 samples were positive for del-HV69/70, but not N501Y, which was confirmed by the TIB MOL reference assays (*table 3*). This is in line with existing data showing sporadic occurrences of del-HV69/70 mutations within the endemic SARS-CoV-2 population (approx. 6%).

**Table 3:**
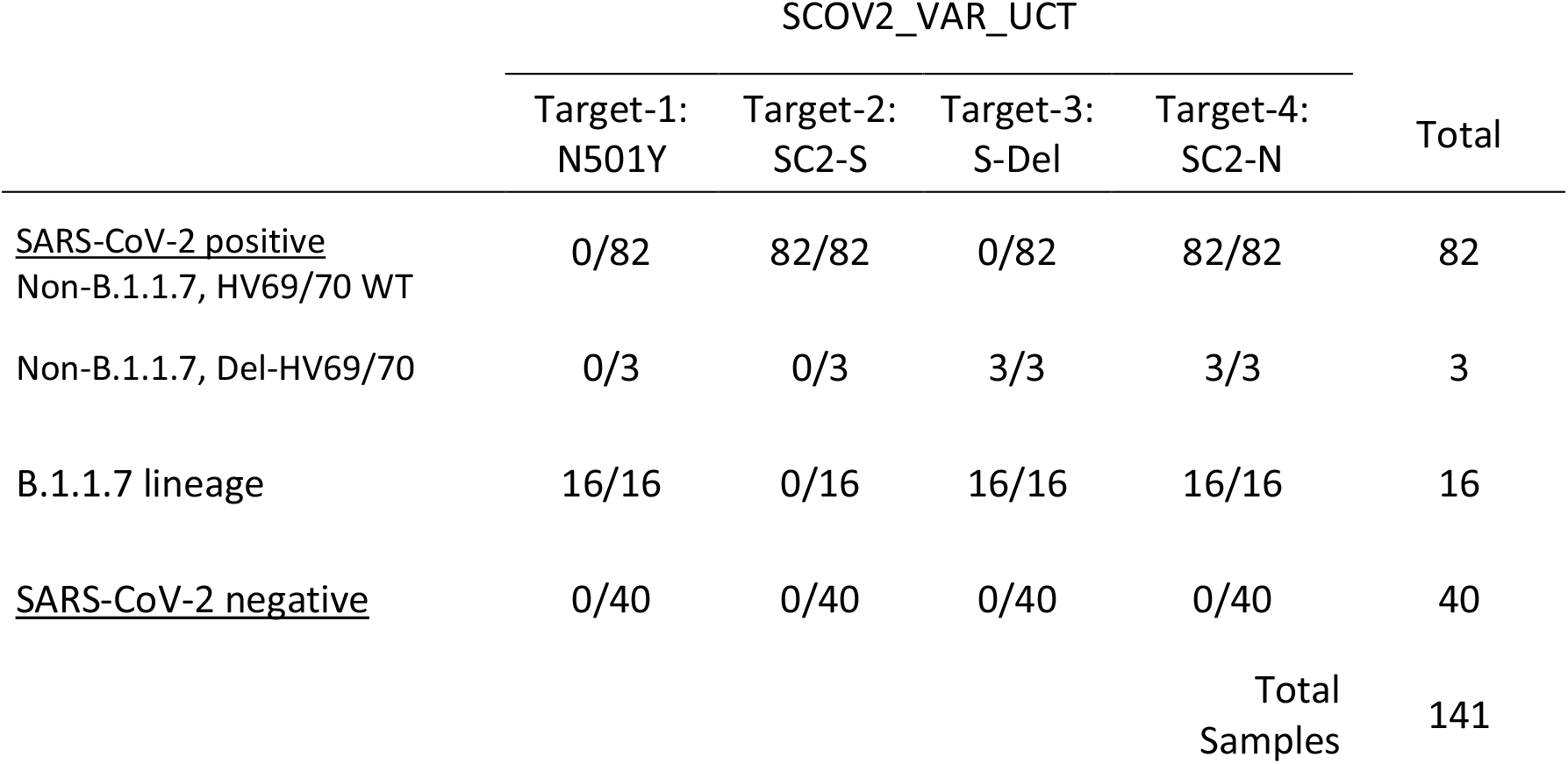
Clinical samples (UTM based) were predetermined positive or negative in routine diagnostics (commercial and inhouse methods) and checked for the Del-69/70 and N501Y spike-gene mutations using commercial VirSNiP assays by TIB MOL (Berlin, Germany). B.1.1.7 lineage samples were predetermined by whole genome sequencing. All samples and mutations were correctly detected by the SCOV2_VAR assay.

## 5 Discussion and Conclusion

Diagnostic labs in Europe are increasingly confronted with demands for rapid differentiation of SARS-CoV-2 isolates due to concerns about potentially higher transmissibility and immune escape with recently emerged lineages such as B.1.1.7 and B1.351 (1, 6). Previous reports have demonstrated how inclusivity issues of commercial assays such as the “TaqPath RT-PCR COVID-19 kit” (Thermo Fischer Scientific, USA) can coincidentally be used to pre-screen for B.1.1.7 lineage variants because of a drop-out phenomenon in one of the targets associated with del-HV69/70 (12). However, this method runs the risk of misinterpreting sporadic HV69/70 deletions, of which we found three within the B.1.1.7 negative set. Mutations like N501Y have independently occurred multiple times in areas with high incidence rates, implying advantages in an environment of high background host-immunity. Their relevance may further grow in the context of ongoing vaccination campaigns. Commercial solutions for SARS-CoV-2 SNP detection such as the TIB MOL VirSNiP-kits are in the process of entering the diagnostics market; however, they mostly consist of manual protocols, thus limiting their suitability for large scale application. The SCOV2_VAR_UCT is able to detect both the del-HV69/70 and N501Y mutations, relevant for B.1.1.7 and B1.351 and further adaptions can be implemented to also pick up new emerging SNPs if necessary. It is self-evident that such methods cannot be a replacement for whole genome sequencing; they do, however, represent a valuable asset to quickly detect clusters and help better direct NGS capacities.

In conclusion, the SCOV2_VAR_UCT multiplex presented in this study combines highly sensitive SARS-CoV-2 detection with relatively reliable identification of the B.1.1.7 lineage by detecting two hallmark mutations. Furthermore, other lineages featuring the N501Y SNP, but lacking the deletion, would also be recognized as abnormal for further investigation. It can thus be used either as a secondary assay for pre-screening prior to sequencing, or as a first-line diagnostic assay. Using the cobas6800/8800 automated systems, this setup can be employed for high-throughput screening for B.1.1.7 and other variants, requiring minimal hands-on time.

## Data Availability

Upon request

## 6 Author contribution

ML, SP, DN, FO, NF and MA conceptualized and supervised the study. DN and MG performed the experiments. DN, ML, SP, NF, FO, and MA wrote and edited the manuscript. All authors agreed to the publication of the final manuscript.

