## Supplementary tables for "Evaluation of a fully automated high-throughput SARS-CoV-2 multiplex qPCR assay with build-in screening functionality for DelHV69/70- and N501Y variants such as B.1.1.7"

### Supplementary table 1

| <u>Cross-reactivity Panel</u><br>Species | - | N501Y | SC2-S | S-DEL | SC2-N |
|---|---|---|---|---|---|
| Influenza A H1N1 pdm09 *(A/California/60/2008)* | | Negative | Negative | Negative | Negative |
| Influenza B Victoria *(B/Brisbane/60/2008)* | | Negative | Negative | Negative | Negative |
| RSV A | | Negative | Negative | Negative | Negative |
| MERS Coronavirus | | Negative | Negative | Negative | Negative |
| hCoV - OC43 | | Negative | Negative | Negative | Negative |
| Human Metapneumovirus (Subtype A) | | Negative | Negative | Negative | Negative |
| Parainfluenzavirus 2 | | Negative | Negative | Negative | Negative |
| Parainfluenzavirus 3 | | Negative | Negative | Negative | Negative |
| Rhinovirus | | Negative | Negative | Negative | Negative |
| hCoV - HKU1 (clinical sample) | | Negative | Negative | Negative | Negative |
| hCoV - NL63 (clinical sample) | | Negative | Negative | Negative | Negative |
| Adenovirus (clinical sample) | | Negative | Negative | Negative | Negative |
| Boca-virus (clinical sample) | | Negative | Negative | Negative | Negative |
| Mycoplasma pneumoniae (clinical sample) | | Negative | Negative | Negative | Negative |
| Pneumocystis jirovecii (clinical sample) | | Negative | Negative | Negative | Negative |

### Supplementary table 2

SARS-CoV-2 "wild-type" (Accuplex Verification Panel)

| Step | cp/ml | SC2-S: pos/rep | SC2-N: pos/rep | Overall: pos/rep |
|---|---|---|---|---|
| 1 | 500.00 | 8/8 | 8/8 | 8/8 |
| 2 | 250.00 | 8/8 | 8/8 | 8/8 |
| 3 | 125.00 | 8/8 | 8/8 | 8/8 |
| 4 | 62.50 | 8/8 | 8/8 | 8/8 |
| 5 | 31.25 | 8/8 | 8/8 | 8/8 |
| 6 | 15.63 | 8/8 | 8/8 | 8/8 |
| 7 | 7.81 | 6/8 | 7/8 | 8/8 |
| 8 | 3.91 | 4/8 | 5/8 | 6/8 |
| 9 | 1.95 | 4/8 | 2/8 | 5/8 |

SARS-CoV-2 B.1.1.7 lineage (clinical sample)

| Step | cp/ml | N501Y: pos/rep | S-Del: pos/rep | SC2-N: pos/rep | Overall: pos/rep |
|---|---|---|---|---|---|
| 1 | 5561.00 | 8/8 | 8/8 | 8/8 | 8/8 |
| 2 | 2780.50 | 8/8 | 8/8 | 8/8 | 8/8 |
| 3 | 1390.25 | 8/8 | 8/8 | 8/8 | 8/8 |
| 4 | 695.13 | 8/8 | 8/8 | 8/8 | 8/8 |
| 5 | 347.56 | 8/8 | 8/8 | 8/8 | 8/8 |
| 6 | 173.78 | 8/8 | 8/8 | 8/8 | 8/8 |
| 7 | 86.89 | 2/8 | 7/8 | 8/8 | 8/8 |
| 8 | 43.45 | 1/8 | 7/8 | 8/8 | 8/8 |
| 9 | 21.72 | 0/8 | 2/8 | 7/8 | 7/8 |
| 10 | 10.86 | 0/8 | 1/8 | 6/8 | 6/8 |
